# Relative contribution of leaving home for work or education, transport, shopping and other activities on risk of acquiring COVID-19 infection outside the household in the second wave of the pandemic in England and Wales

**DOI:** 10.1101/2021.12.08.21267458

**Authors:** S Hoskins, S Beale, RW Aldridge, AMD Navaratnam, C Smith, CE French, A Yavlinsky, V Nguyen, T Byrne, J Kovar, E Fragaszy, WLA Fong, C Geismar, P Patel, AM Johnson, Hayward AC - on behalf of the Virus Watch Collaborative

## Abstract

**Background:** With the potential for and emergence of new COVID-19 variants, such as the reportedly more infectious Omicron, and their potential to escape the existing vaccines, understanding the relative importance of which non-household activities increase risk of acquisition of COVID-19 infection is vital to inform mitigation strategies.

**Methods:** Within an adult subset of the Virus Watch community cohort study, we sought to identify which non-household activities increased risk of acquisition of COVID-19 infection and which accounted for the greatest proportion of non-household acquired COVID-19 infections during the second wave of the pandemic. Among participants who were undertaking antibody tests and self-reporting PCR and lateral flow tests taken through the national testing programme, we identified those who were thought to be infected outside the household during the second wave of the pandemic. We used exposure data on attending work, using public or shared transport, using shops and other non-household activities taken from monthly surveys during the second wave of the pandemic. We used multivariable logistic regression models to assess the relative independent contribution of these exposures on risk of acquiring infection outside the household. We calculated Adjusted Population Attributable Fractions (APAF - the proportion of non-household transmission in the cohort thought to be attributable to each exposure) based on odds ratios and frequency of exposure in cases.

**Results:** Based on analysis of 10475 adult participants including 874 infections acquired outside the household, infection was independently associated with: leaving home for work (AOR 1.20 (1.02 – 1.42) p=0.0307, APAF 6.9%); public transport use (AOR for use more than once per week 1.82 (1.49 – 2.23) p<0.0001, APAF for public transport 12.42%); and shopping (AOR for shopping more than once per week 1.69 (1.29 – 2.21) P=0.0003, APAF for shopping 34.56%). Other non-household activities such as use of hospitality and leisure venues were rare due to restrictions and there were no significant associations with infection risk.

**Conclusions:** A high proportion of the second wave of the pandemic was spent under conditions where people were being advised to work from home where possible, and to minimize exposure to shops, and a wide range of other businesses were subject to severe restrictions. Vaccines were being rolled out to high-risk groups. During this time, going to work was an important risk factor for infection but public transport use likely accounted for a lot of this risk. Only a minority of the cohort left home for work or used public or shared transport. By contrast, the majority of participants visited shops and this activity accounted for about one-third of non-household transmission.

## Introduction

The COVID-19 pandemic has led to levels of hospitalisation and mortality that are unprecedented in recent history. Governments around the world have imposed strict impositions on social mixing to control the virus. The relative importance of these restrictions is challenging to estimate as empirical evidence to date relies on ecological studies where it can be difficult to disentangle the effects of multiple interventions introduced at different stages of the pandemic in different countries and regions. A review of such empirical studies from the first wave of the pandemic suggests that school and workplace closures, closure of businesses and banning of public events had the greatest impact on transmission, especially when introduced early. (1)

The effectiveness of restrictions depends on the amount of transmission that occurs in different settings and whilst transmission can take place in any setting the relative importance of different non-household settings and activities to spread of infection is poorly understood. (2)

Occupation is known to be an important predictor of COVID-19 infection and mortality risk, with a high proportion of the differential risk between occupations likely to be related to differential ability to work from home during periods of intense COVID-19 transmission. (3)

Transmission on public transport has also been shown to occur but the relative importance of this compared to other exposures is unknown and since transport is used as a means of travelling to undertake other activities, it has proved challenging to untangle the risk from that of other out of home exposures. (4)

Although it is clear that the importance of different venues such as hospitality, retail and leisure on population infection rates depends on both the likelihood of transmission occurring within a particular environment and the frequency with which people visit that setting, the relative importance of these settings has proved difficult to assess.(5) During periods of intense control essential shops have remained open but the amount of transmission occurring in such settings is unknown.

We aimed to understand the relative importance of different activities and settings in the transmission of COVID-19 in England and Wales during the second wave of the pandemic, a period with a relatively intensive range of non-pharmaceutical interventions (NPI’s) including advice to work from home where possible, closure of a range of non-essential businesses such as hospitality and leisure venues and restrictions on social gatherings.

## Methods

The analyses are based on the Virus Watch Cohort, the detailed methodology of this cohort is described elsewhere (6). Briefly, the study recruits whole households with detailed baseline information, weekly surveys of symptoms and self-reporting of positive COVID-19 tests (PCR or lateral flow) conducted through the national tracing programme, linkage to the national testing data set, and monthly questionnaires on contact and activity patterns. A subset of the adult cohort have also undertaken antibody testing through venous blood draws since October 2020 and monthly finger-prick testing since March 2021.

Within an adult subset of the Virus Watch community cohort study, we identified a subcohort of participants who completed two monthly behavioural surveys during the second wave of the pandemic (completed during the periods 1/12/2020 – 10/12/2020 and 17/02/2021 – 28/02/2021). These asked about the frequency of going to work, using public transport, going to retail settings and visiting other non-household settings in the week prior to the survey. These were averaged to give the average weekly frequency of these activities. We also asked about the number of known close contacts outside the household during these weeks, where close contact was defined as being within 2m of someone for more than 15 minutes.

We further restricted the cohort to those who were taking part in the antibody test study. Anyone who tested positive for nucleocapsid antibody due to likely infection versus vaccination between 01/10/2020 and 01/05/2021 was considered to have been infected in the second wave of the pandemic unless they had previously reported a positive PCR or lateral flow test prior to the second wave. Participants who tested positive for antibodies on venous blood samples taken before the second wave were excluded. Anyone testing PCR or lateral flow positive during the second wave (01/10/2020 - 01/05/2021) was also considered to have been infected during the second wave.

In order to focus analyses on risk factors for non-household transmission we further restricted cases to those which were the only case in the household (no other antibody or PCR or lateral flow cases in the household) and, in households with more than one PCR/lateral flow confirmed case, to i) the first case in the household based on the earliest date of symptom onset in the house, or ii) where there were multiple positive people in the household reporting the same symptom onset date, all cases were included as co-primaries, or iii) if no symptom data were available, based on the earliest date of PCR or lateral flow test in the house using dates from the National Testing data, or if National testing date data were not available, using the Virus Watch given date or imputed middle of the week date when the test was self-reported. Where we could not identify who was the likely first case in the household these infections were excluded from the analyses.

We undertook univariate analyses comparing the proportion with evidence of infection acquired outside the household, according to different weekly frequency of going to work, using public or shared transport, using retail venues and other non-household contacts. We conducted multivariate logistic regression models including these variables as well as age, gender, vaccination status, social deprivation, region and ethnicity. For activities associated with risk of non-household transmission we calculated stratum specific univariate and multivariate population attributable fractions during the second wave of the pandemic. We used the formula PAF = p * (1-1/Relative Risk) where p=proportion of those with COVID-19 acquired outside the household who had the exposure of interest and where adjusted and unadjusted odds ratios were taken as proxies of relative risk (based on the rare outcome assumption).

## Results

Of the 10,858 Virus Watch participants who completed the two monthly surveys and were members of the antibody cohort, 1,257 (12%) were identified as having had Covid-19 (343 were identified by either PCR or lateral flow test and 914 by likely infection-induced antibody results alone). Of all Covid-19 cases identified via PCR or lateral flow test, we estimate up to 18% of these may be due to household transmission based on the timing of illnesses and swabs in other household members (Table 1).

**Table 1:** Proportion of overall infection attributable to household and non-household transmission

| **Identification of primary, co-primary and secondary cases** | **Full monthly survey AB cohort (n=10,858)** |
| --- | --- |
| Number of positive PCR/lat flow | 343 |
| b) Secondary cases (no symptom data but someone in the house tests (+) first, by National Testing. If national testing data date was not available then the date of onset was imputed as the middle of the week when the participant reported they tested positive). | 16(5%) |
| a)Secondary case (someone else in home who is PCR/Lateral Flow positive has symptoms prior to case) | 47 (14%) |
| Upper Limit Household transmission (proportion of PCR confirmed cases that are thought to be secondary cases) (a+b) | 63/343=18% |
| c)Primary case: Only case in a home | 189 (55%) |
| d) Co-primary case: Symptom date the same as earliest symptom in the home | 85 (25%) |
| e) Primary case: no symptom data but tests first in the home by National Testing data. If National testing data not available, by VW imputed mid-week date. | 6 (2%) |
| Non-household transmission - proportion of PCR/lateral flow confirmed cases that are thought to be due to non-household transmission (c+d+e/a+b+c+d+e) | 280/343=82% |
| AB (+) PCR (-) cases | 914 |
| e) Primary AB case: only AB (+) PCR (-) case in the home (i.e. no other AB (+) or PCR (+) case in the home) | 594 (65%) |
| f) Potential secondary case: AB (+) PCR (-) case with someone else infected in the home either as<br>- another AB (+) PCR (-) case in the home or<br>- a (+) PCR case in the home | 279<br>41<br>Total 320 (35%) |
| Non-household transmission cohort = 10858-(a+b+f) | 10,475 |

The non-household transmission study therefore included 10475 participants; 874 COVID-19 cases identified through PCR, lateral flow and/or positive antibody tests who were thought to have been infected outside the household during the second wave of the pandemic and 9601 uninfected participants (Table 1). The breakdown of the cohort by age, gender, index of multiple deprivation, region, ethnicity and vaccination status are shown in table 2 with univariate analyses of the risk of infection acquired outside the household in each group. The highest levels of infection were seen in working-aged adults, women, poorer areas, non-white ethnic groups, London and the unvaccinated.

**Table 2.** Participant characteristics and risk of infection acquired outside the household during the pandemic second wave

| Characteristic | Category | N=10,475<br>(% in<br>category) | Number of<br>infections<br>acquired<br>outside the<br>household<br>n=874 (%<br>within<br>category) | Unadjusted<br>OR | 95% CI | P |
| --- | --- | --- | --- | --- | --- | --- |
| Vaccine status | Yes<br>No | 9,502 (91%)<br>973 (9%) | 736 (8%)<br>138 (14%) | 1.00<br>1.97 | 1.62 – 2.39 | <0.0001 |
| Age | Under 18<br>Working Age<br>65 and above<br>Missing | 383 (4%)<br>5,412 (52%)<br>4,669 (45%)<br>11 | 23 (6%)<br>634 (12%)<br>217 (5%) | 1.00<br>2.08<br>0.76 | 1.35 – 3.19<br>0.49 – 1.19 | <0.0001 |
| Sex | Male<br>Female<br>Missing | 4,538 (43%)<br>5,913 (57%)<br>24 | 308 (7%)<br>563 (10%) | 1.00<br>1.44 | 1.25 – 1.67 | <0.0001 |
| Ethnic group | White<br>White Other<br>Asian<br>Black<br>Mixed<br>Other<br>Missing | 9,475 (91%)<br>589 (6%)<br>240 (2%)<br>55 (1%)<br>59 (1%)<br>49 (<1%)<br>8 | 774 (8%)<br>54 (9%)<br>31 (13%)<br>5 (9%)<br>6 (10%)<br>4 (8%) | 1.00<br>1.13<br>1.67<br>1.12<br>1.27<br>0.99 | 0.85 – 1.52<br>1.14 – 2.45<br>0.45 – 2.83<br>0.55 – 2.97<br>0.36 – 2.79 | 0.2302 |
| Deprivation score<br>(IMD quintile) 1=<br>most deprived | 1<br>2<br>3<br>4<br>5<br>Missing | 778 (7%)<br>1,538 (15%)<br>2,099 (20%)<br>2,815 (27%)<br>3,230 (31%)<br>15 | 71 (9%)<br>165 (11%)<br>177 (8%)<br>225 (8%)<br>236 (7%) | 1.27<br>1.52<br>1.17<br>1.10<br>1.00 | 0.97 – 1.68<br>1.24 – 1.88<br>0.95 – 1.43<br>0.91 – 1.33 | 0.0026 |
| Region | East Midlands | 1,067 (10%) | 83 (8%) | 1.00 |  | <0.0001 |
|  | E.of England | 2,236 (21%) | 160 (7%) | 0.91 | 0.69 – 1.20 |  |
|  | London | 1,125 (11%) | 159 (14%) | 1.95 | 1.48 – 2.58 |  |
|  | North East | 554 (5%) | 51 (9%) | 1.20 | 0.83 – 1.73 |  |
|  | North West | 1,233 (12%) | 96 (8%) | 1.00 | 0.74 – 1.36 |  |
|  | South East | 1,840 (18%) | 146 (8%) | 1.02 | 0.77 – 1.35 |  |
|  | South West | 948 (9%) | 54 (6%) | 0.72 | 0.50 – 1.02 |  |
|  | Wales | 255 (2%) | 24 (9%) | 1.23 | 0.76 – 1.98 |  |
|  | W. Midlands | 588 (6%) | 49 (8%) | 1.08 | 0.75 – 1.56 |  |
|  | Yorkshire &<br>The Humber | 614 (6%) | 52 (8%) | 1.09 | 0.76 – 1.58 |  |
|  | Missing | 15 |  |  |  |  |

Supplementary table S1 shows the detailed breakdown of public or shared transport exposures in relation to risk of infection. On univariate analysis, all forms of public or shared transport, with the exception of using an airplane which was rare, were associated with an increased risk of COVID-19 infection. Table S2 shows the detailed breakdown of non-work, non-public or shared transport use and out of household activities and associations with non-household acquired infections. The only significant association during this period was with essential retail, although exposure to other settings such as leisure and hospitality was rare during this period.

Table 3 shows the relative impact of leaving home to go to work or education, using public or shared transport, retail and other non-household activities on the risk of infection acquired outside the household. Models are adjusted for all variables in the table in addition to age, gender, ethnicity, region, vaccination and index of multiple deprivation quintile. Table 3 also shows the unadjusted and adjusted population attributable fractions for leaving home for work, public or shared transport and retail.

**Table 3.** Unadjusted and adjusted odds ratios and population attributable fractions for non-household COVID-19 acquisition

| Activity and frequency of occurrence |  |  | Positive PCR lateral flow or AB | Univariate and multivariate analyses |  | Population attributable fraction |
| --- | --- | --- | --- | --- | --- | --- |
| Activity | Weekly frequency | Total in AB cohort (n=10,475) | Number of COVID cases (n=874) | OR (95% CI) ,p | Adjusted OR (95% CI), p | (%) Raw (adjusted) |
| Leaving home for work or education | No<br>Yes | 7,266 (69%)<br>3,209 (31%) | 507 (7%)<br>367 (11%) | 1<br>1.72 (1.49 – 1.98)<br>p<0.001 | 1.00<br>1.20 (1.02 – 1.42)<br>P=0.0307 | 17.58 (Adj-6.99) |
| Weekly frequency of using public transport | 0<br>>0 -1<br>>1 | 7,621 (73%)<br>1,733 (17%)<br>1,121 (11%) | 539 (7%)<br>165 (10%)<br>170 (15%) | 1.00<br>1.38 (1.15 – 1.66)<br>2.35 (1.95 – 2.83)<br>P<0.001 | 1.00<br>1.24 (1.03 – 1.49)<br>1.82 (1.49 – 2.23)<br>p<0.0001 | 5.19 (Adj-3.65)<br>11.17(Adj-8.76)<br>Total<br>16.37 (Adj 12.42) |
| Weekly frequency of any retail | 0<br>>0 -1<br>>1 | 1,560 (15%)<br>3,030 (29%)<br>5,885 (56%) | 78 (5%)<br>234 (8%)<br>562 (10%) | 1.00<br>1.59 (1.22 – 2.07)<br>2.01 (1.57 – 2.56)<br>P<0.001 | 1.00<br>1.45 (1.09 – 1.92)<br>1.69 (1.29 – 2.21)<br>P=0.0003 | 9.93 (Adj-8.31)<br>32.31 (Adj-26.25)<br>Total<br>42.25 (Adj 34.56) |
| Weekly frequency of other non household activities | 0 | 823 (8%) | 51 (6%) | 1.00 | 1.00 |  |
|  | >0 – <2 | 2,454 (23%) | 184 (8%) | 1.23 (0.89 – 1.69) | 0.99 (0.71 – 1.39) | 3.94 |
|  | 2 – 3 | 2,396 (23%) | 191 (8%) | 1.31 (0.95 – 1.81) | 0.88 (0.63 – 1.23) | 5.17 |
|  | >3 – 5 | 2,620 (25%) | 248 (9%) | 1.58 (1.16 – 2.16) | 0.97 (0.69 – 1.36) | 10.41 |
|  | > 5 | 2,182 (21%) | 200 (9%) | 1.53 (1.11 – 2.10) | 0.87 (0.61 – 1.23) | 7.93 |
|  |  |  |  | P<0.0066 | P=0.6188 | Total 27.45<br>(adjusted not estimated) |

The odds ratio for leaving home for work or education at least once per week was 1.72 but this association was considerably weakened after controlling for public transport use and other risk factors - adjusted odds ratio 1.20 (1.02 – 1.42) p=0.0307. Across the cohort leaving home for work accounted for 18% of non-household acquired infections (PAF) but this was reduced to 7% after controlling for transport use and other variables.

The odds ratio for using public or shared transport more than once per week compared to no usage was 2.35 but this was significantly reduced after controlling for going to work or education and other risk factors (adjusted OR 1.82 (1.49 – 2.23) p<0.0001). Across the cohort public or shared transport use accounted for 16% of infections acquired outside the household, though this was reduced to 12% after controlling for going to work and other variables.

The odds ratio for using shops more than once per week compared to no usage was 2.01 but reduced after controlling for other variables (adjusted OR 1.69 (1.29 – 2.21) P=0.0003). Across the cohort, shopping accounted for 42% of infections acquired outside the household and 35% after controlling for other variables.

Notably, despite significant univariate associations between ethnicity and social deprivation and COVID risk, the full final multivariate model (table S5) including work, public or shared transport use, shopping and other variables showed no significant difference in risk of acquiring COVID-19 outside the household by ethnicity (p= 0.12) or social deprivation quintile (p=0.71).

Supplementary tables S3 and S4 show the unadjusted and adjusted odds ratios and PAFs for infection acquired outside the household in the working-age population (18-64) and in those aged > 64 respectively. Of note, among those of working age, leaving home for work or education and using public or shared transport each accounted for around 10% of non-household acquired infections (aPAFs 8.99% and 10.94%, respectively) while using shops accounted for 32% (aPAF 31.87%). For those aged 65 years and above, using public or shared transport (aPAF 14.06%) and shopping (aPAF 38.41%) accounted for the greatest proportion of non-household acquired infections. Leaving home for work was rare in this age group and not significantly associated with risk of infection.

Supplementary table S5 shows the effect of leaving home for work, public or shared transport use and shopping after additionally controlling for the number of close contacts outside the home and the previously described adjustment variables. The remaining risk from attending work is further ameliorated (aOR 1.10 95% CI 0.92 – 1.32 p= 0.30). Adjusted odds ratios for public transport and shopping were minimally affected by adding the number of non-household close contacts as a covariate.

## Discussion

The study demonstrates that leaving home for work or education, public transport and shopping were important independent risk factors for acquiring COVID-19 outside the household during the second wave of the pandemic in England and Wales. Although those going to work had a substantially higher risk of infection much of this was explained by public or shared transport use and other variables. This suggests that a high proportion of the risk associated with going to work or education was due to exposure on public or shared transport. Public or shared transport use remained a strong independent risk factor after controlling for other variables. Shopping was also an important risk factor for acquiring COVID-19 outside the household and, because it was a very common exposure, accounted for a high proportion of infections acquired outside the household in adults. Other non-household activities such as visiting hospitality or leisure venues were not significantly associated with acquiring COVID-19 outside the household but were rare during this period of intense restrictions. The risk of infection acquired outside the household was strongly associated with the number of close contacts outside the household. Controlling for this further ameliorated the effect of attending work suggesting this is mediated by close contact at work. Controlling for close contact made little difference to associations with public or shared transport and shopping suggesting these exposures are not mediated by recognized close contact and may represent more distant aerosol-based transmission (7). It was interesting that the effect of ethnicity and social deprivation was not seen after accounting for work, transport use, shopping and other variables – suggesting different patterns of exposure to work, associated public or shared transport use and use of shops may account for differential infection rates.

By restricting our analysis to those in the cohort with antibody test results we could ensure that infection could be ascertained in all participants (although antibody waning may lead to loss of detectable nucleocapsid antibody). Due to limited testing capacity nationwide during the first wave of the pandemic, it is possible that we have included some individuals with positive antibodies as a result of infection prior to October 2020. By excluding those with positive PCR or lateral flow or antibody tests prior to October 2020, we sought to minimise this potential over-ascertainment bias. While we sought to identify cases who were the first or only case of COVID-19 in the household it is possible that there was some misclassification error because of uncertainties in the timing of infection and failure to identify all infections in a household. Activities and behaviours are self-reported and therefore subject to recall bias, we tried to minimise this by asking about activities in the previous seven days. These activities were sampled at two points during the second wave and may not be representative of the activities throughout the second wave.

Both self-reported and linked data on test results from the national testing system also allowed ascertainment of infections. Maximising ascertainment of COVID-19 infections supports accurate assessment of the relative importance of risk factors. A further strength was the household structure of the cohort allowing us to focus analyses onto risk factors for non-household transmission and largely eliminate confounding effects that act on household transmission, although misclassification will have occurred. Outcomes and exposures were both measured during the same wave of the pandemic, although for those with antibody results only it was not possible to ascertain whether they were infected during the first or the second wave of the pandemic. Population attributable fractions are influenced by the frequency of exposures within the cohort which may not be representative of the entire adult population. For example, if our cohort includes fewer people going to work than on average, then this will lead to underestimation of the PAF related to going to work.

The research suggests that working from home and consequently avoiding the need to use public or shared transport has a significant impact on risk at individual and population level. We could not ascertain the risk associated with transmission in hospitality and leisure venues as such exposures were rare, suggesting that regulations restricting their use was effective in reducing transmission. Shopping which remained a common exposure was an important contributor to risk at individual and population levels.

During periods of intense COVID-19 transmission, increasing the proportion of people who work from home, facilitating active transport such as cycling or walking in those who need to go to work, and enabling people to shop for essential goods online would be expected to make a highly significant impact on transmission and risk of severe disease. Although high vaccination rates reduce the need for intense non pharmaceutical interventions these measures remain important in poorly vaccinated countries and may become important in the event of COVID-19 resurgence due to waning immunity, increased population mixing or emergence of new variants such as the Omicron variant.

## Data Availability

Data availability
We aim to share aggregate data from this project on our website and via a "Findings so far" section on our website - https://ucl-virus-watch.net/. We will also be sharing individual record level data on the Office of National Statistics Secure Research Service. In sharing the data we will work within the principles set out in the UKRI Guidance on best practice in the management of research data. Access to use of the data whilst research is being conducted will be managed by the Chief Investigators (ACH and RWA) in accordance with the principles set out in the UKRI guidance on best practice in the management of research data. We will put analysis code on publicly available repositories to enable their reuse.

## Ethics

The Virus Watch study has been approved by the Hampstead NHS Health Research Authority Ethics Committee. Ethics approval number - 20/HRA/2320.

## Data availability

We aim to share aggregate data from this project on our website and via a “Findings so far” section on our website - https://ucl-virus-watch.net/. We will also be sharing individual record level data on the Office of National Statistics Secure Research Service. In sharing the data we will work within the principles set out in the UKRI Guidance on best practice in the management of research data. Access to use of the data whilst research is being conducted will be managed by the Chief Investigators (ACH and RWA) in accordance with the principles set out in the UKRI guidance on best practice in the management of research data. We will put analysis code on publicly available repositories to enable their reuse.

**Virus Watch Collaborative:

Susan Michie, Pia Hardelid, Linda Wijlaars, Eleni Nastouli, Moira Spyer, Ben Killingley, Ingemar Cox, Vasileios Lampos, Rachel A McKendry, Tao Cheng, Yunzhe Liu, Jo Gibbs, Richard Gilson, Anne M Johnson, Alison Rodger

Centre for Behaviour Change, University College London, London, UK (SM); Department of Population, Policy and Practice, UCL Great Ormond Street Institute of Child Health, London, UK (PH, LW, EN, MS, BK); Francis Crick Institute, London, UK (EN, MS); Health Protection and Influenza Research Group, Division of Epidemiology and Public Health, University of Nottingham School of Medicine, Nottingham, UK (BK); University College London Hospital, London, UK (BK); Department of Computer Science, University College London, London, UK (IC, VL); London Centre for Nanotechnology and Division of Medicine, London, University College London (RM); SpaceTimeLab, Department of Civil, Environmental and Geomatic Engineering, University College London, London, UK (TC, YL); Institute for Global Health, University College London, London, UK (JG, RG). Institute for Global Health, University College London, London, UK (AMJ, AR).

## Supplementary materials

**Table S1.**
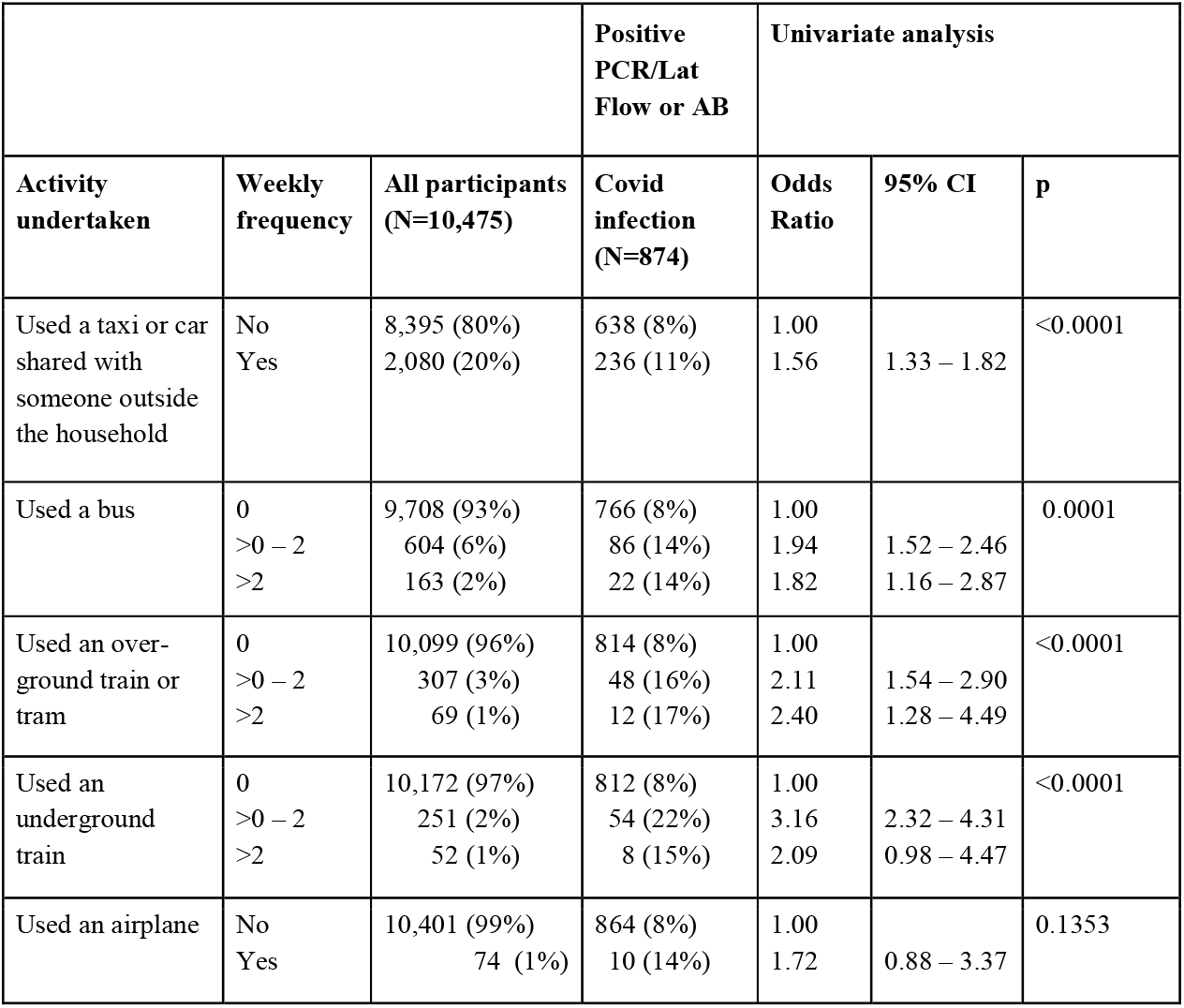
Risk of infection according to type and frequency of public or shared transport use

**Table S2.** Risk of infection according to frequency of non-work or education and non-public or shared transport activities outside the household

|  |  |  | Positive<br>PCR/Lat<br>Flow or AB | Univariate analysis |  |  |
| --- | --- | --- | --- | --- | --- | --- |
| Activities | Weekly<br>frequency | All participants<br>N=10,475 | Covid<br>infection<br>N=874 | Odds<br>Ratio | 95% CI | p |
| Played a team<br>sport outdoors | No<br>Yes | 10,319 (99%)<br>156 (1%) | 862 (8%)<br>12 (8%) | 1.00<br>0.91 | 0.51– 1.65 | 0.7642 |
| Went to a theatre,<br>cinema, concert or<br>sports event | No<br>Yes | 10,430 (99%)<br>45 (<1%) | 868 (8%)<br>6 (13%) | 1.00<br>1.69 | 0.72 – 4.01 | 0.2602 |
| Went to a shop for<br>essential items | 0<br>>0 – 2<br>>2 – 4<br>>4 – 5<br>>5 | 1,748 (17%)<br>6,273 (60%)<br>2,063 (20%)<br>219 (2%)<br>172 (2%) | 90 (5%)<br>541 (9%)<br>202 (10%)<br>24 (11%)<br>17 (10%) | 1.00<br>1.74<br>1.99<br>2.27<br>2.02 | 1.38 – 2.19<br>1.55 – 2.59<br>1.41 – 3.64<br>1.17 – 3.48 | <0.0001 |
| Went to a shop for<br>non-essential<br>items | No<br>Yes | 8,187 (78%)<br>2,288 (22%) | 677 (8%)<br>197 (9%) | 1.00<br>1.05 | 0.89 – 1.23 | 0.6034 |
| Went to a bar,<br>pub, club, disco | No<br>Yes | 10,361 (99%)<br>114 (1%) | 864 (8%)<br>10 (9%) | 1.00<br>1.06 | 0.55 – 2.03 | 0.8689 |
| Ate at a restaurant,<br>café or canteen | No<br>Yes | 10,100 (96%)<br>375 (4%) | 834 (8%)<br>40 (11%) | 1.00<br>1.33 | 0.95 – 1.86 | 0.1104 |
| Went to a party | No<br>Yes | 10,446 (99%)<br>29 (<1%) | 872 (8%)<br>2 (7%) | 1.00<br>0.81 | 0.19 – 3.43 | 0.7717 |
| Went to a place of<br>worship | No<br>Yes | 10,124 (97%)<br>351 (3%) | 849 (8%)<br>25 (7%) | 1.00<br>0.84 | 0.55 – 1.27 | 0.3894 |
| Went to an<br>outdoor market | No<br>Yes | 9,806 (94%)<br>669 (6%) | 816 (8%)<br>58 (9%) | 1.00<br>1.04 | 0.79 – 1.38 | 0.7540 |
| Went to a<br>gym/indoor sport | No<br>Yes | 10,284 (98%)<br>191 (2%) | 858 (8%)<br>16 (8%) | 1.00<br>1.00 | 0.59 – 1.68 | 0.9866 |
| Went to a<br>hairdresser,<br>barber, nail salon,<br>beauty parlour | No<br>Yes | 10,212 (97%)<br>263 (3%) | 853 (8%)<br>21 (8%) | 1.00<br>0.95 | 0.61 – 1.49 | 0.8301 |

**Table S3.** Unadjusted and adjusted odds ratios and population attributable fractions for non-household COVID-19 acquisition among those of working age (18 - 64 years)

| Activity and frequency of occurrence |  |  | Positive PCR lateral flow or AB | Univariate and multivariate analyses |  | Population Attributable Fraction (%) |
| --- | --- | --- | --- | --- | --- | --- |
| Activity | Weekly frequency | Total in AB cohort (n=5,412) | Number of COVID cases (n=634) | OR (95% CI), p | Adjusted OR (95% CI), p | Raw (adjusted) |
| Leaving home for work or education | No<br>Yes | 3,008(56%)<br>2,404 (44%) | 318 (11%)<br>316 (13%) | 1<br>1.28 (1.08 – 1.51)<br>P=0.0035 | 1.00<br>1.22 (1.02 – 1.47)<br>P=0.0329 | 10.90 (Adj 8.99) |
| Weekly frequency of using public transport | 0<br>>0 -1<br>>1 | 3,850 (71%)<br>900 (17%)<br>662 (12%) | 396 (10%)<br>117 (13%)<br>121 (18%) | 1.00<br>1.30 (1.04 – 1.62)<br>1.95 (1.56 – 2.44)<br>P<0.001 | 1.00<br>1.19 (0.96 – 1.50)<br>1.72 (1.35 – 2.19)<br>P=0.0001 | 4.26 (Adj 2.95)<br>9.29 (Adj 7.99)<br>Total<br>13.56 (Adj 10.94) |
| Weekly frequency<br>of any retail | 0 | 636 (12%) | 50 (8%) | 1.00 | 1.00 |  |
|  | >0 - 1 | 1,540 (28%) | 169 (11%) | 1.44 (1.04 – 2.01) | 1.38 (0.99– 1.94) | 8.14 (Adj 7.34) |
|  | >1 | 3,236 (60%) | 415 (13%) | 1.72 (1.27 – 2.34) | 1.60 (1.16 – 2.22) | 27.4 (Adj 24.55) |
|  |  |  |  | P=0.0006 | P=0.0097 | Total<br>35.55 (Adj 31.87) |
| Weekly frequency<br>of other non<br>household<br>activities | 0 | 349 (6%) | 37 (11%) | 1.00 | 1.00 |  |
|  | >0 – <2 | 1,073 (20%) | 125 (12%) | 1.11 (0.75 – 1.64) | 0.91 (0.61 – 1.35) | 1.95 |
|  | 2 – 3 | 1,210 (22%) | 136 (11%) | 1.07 (0.73 – 1.57) | 0.78 (0.52 – 1.16) | 1.40 |
|  | >3 – 5 | 1,457 (27%) | 171 (12%) | 1.12 (0.77 – 1.63) | 0.77 (0.51 – 1.15) | 2.89 |
|  | > 5 | 1,323 (24%) | 165 (12%) | 1.20 (0.82 – 1.75) | 0.79 (0.53 – 1.21) | 4.34 |
|  |  |  |  | P=0.8404 | P=0.5716 | Total 10.58<br>Adjusted not estimated |

**Table S4.** Unadjusted and adjusted odds ratios and population attributable fractions for non-household COVID-19 acquisition among those of retired age (65 years and above)

| Activity and frequency of occurrence |  |  | Positive PCR lateral flow or AB | Univariate and multivariate analyses |  | Population Attributable Fraction (%) |
| --- | --- | --- | --- | --- | --- | --- |
| Activity | Weekly frequency | Total in AB cohort (n=4,669) | Number of COVID cases (n=217) | OR (95% CI) ,p | Adjusted OR (95% CI), p | Raw (adjusted) |
| Leaving home for work or education | No<br>Yes | 4,220(90%)<br>449 (10%) | 189 (4%)<br>28 (6%) | 1<br>1.42 (0.94 – 2.14)<br>P=0.1073 | 1.00<br>1.28 (0.82 – 1.99)<br>P=0.2806 | 3.82 (Adj 2.82) |
| Weekly frequency of using public transport | 0<br>>0 -1<br>>1 | 3,515(75%)<br>795(17%)<br>359 (8%) | 132 (4%)<br>46 (6%)<br>39 (11%) | 1.00<br>1.57 (1.11 – 2.22)<br>3.12 (2.15 – 4.54)<br>P<0.001 | 1.00<br>1.29 (0.89 – 1.87)<br>2.07 (1.36 – 3.14)<br>P=0.0042 | 7.69 (Adj 4.77)<br>12.21 (Adj 9.29)<br>Total<br>19.91 (Adj 14.06) |
| Weekly frequency of any retail | 0<br>>0 -1<br>>1 | 669 (14%)<br>1,400 (30%)<br>2,600 (56%) | 17 (3%)<br>57 (4%)<br>143 (6%) | 1.00<br>1.62 (0.94 – 2.82)<br>2.23 (1.34 – 3.72)<br>P=0.0014 | 1.00<br>1.59 (0.89 – 2.83)<br>1.77 (1.01 – 3.09)<br>P=0.1083 | 10.05 (Adj 9.75)<br>36.35 (Adj 28.67)<br>Total<br>46.40 (Adj 38.41) |
| Weekly frequency<br>of other non<br>household<br>activities | 0 | 434 (9%) | 14 (3%) | 1.00 | 1.00 |  |
|  | >0 – <2 | 1,340 (29%) | 57 (4%) | 1.33 (0.74 – 2.42) | 1.12 (0.59 – 2.12) | 6.52 (Adj 2.81) |
|  | 2 – 3 | 1,050 (22%) | 49 (5%) | 1.47 (0.80 – 2.69) | 1.07 (0.55 – 2.07) | 7.22 (Adj 1.48) |
|  | >3 – 5 | 1,048 (22%) | 66 (6%) | 2.02 (1.12 – 3.63) | 1.47 (0.77 – 2.80) | 15.36 (Adj 9.72) |
|  | > 5 | 797 (17%) | 31 (4%) | 1.21 (0.64 – 2.31)<br>P=0.0477 | 0.88 (0.43 – 1.78)<br>P=0.2195 | 2.48 (adj not<br>estimated)<br>Total 31.57 (Adj<br>not estimated) |

**Table S5.**
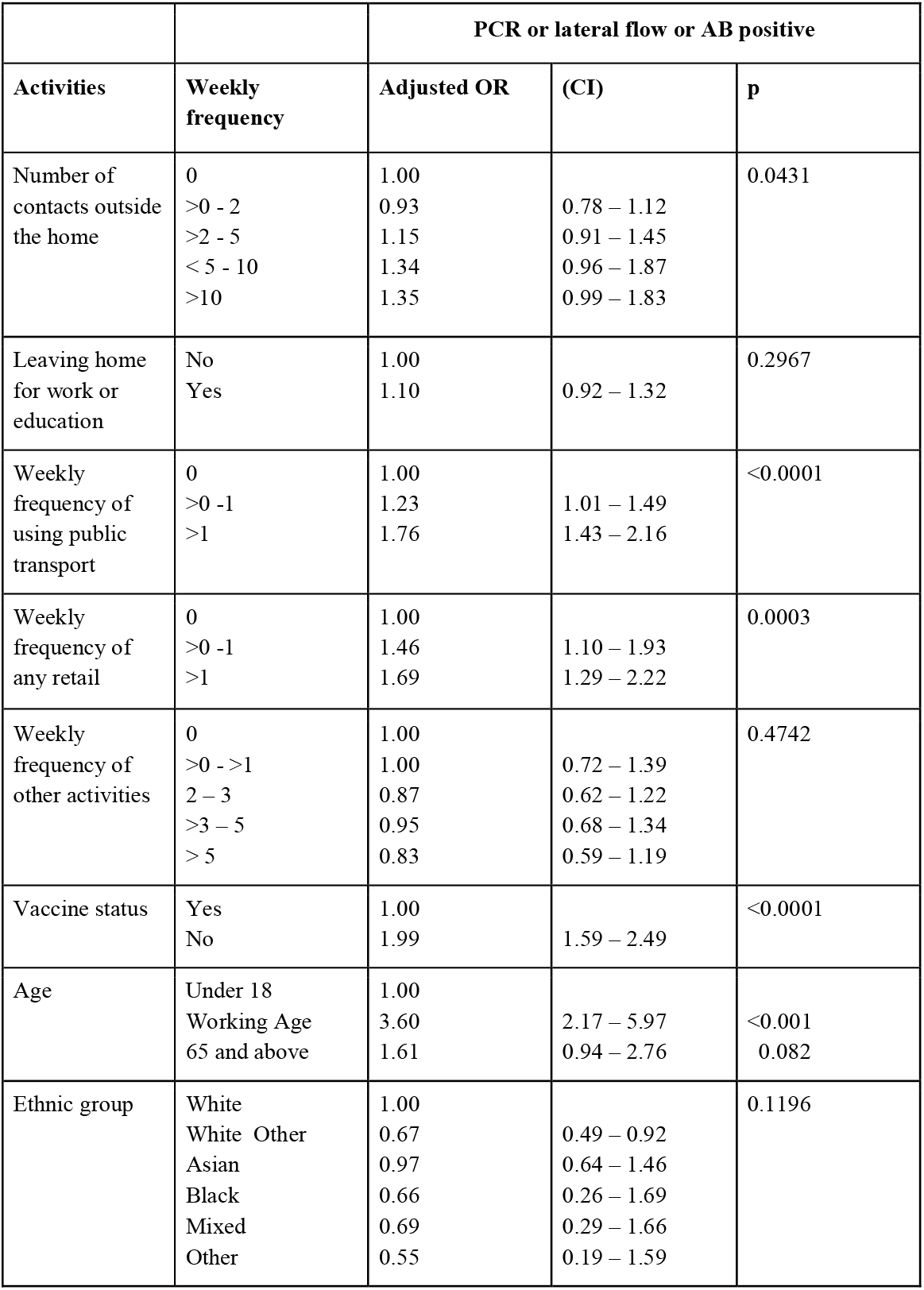
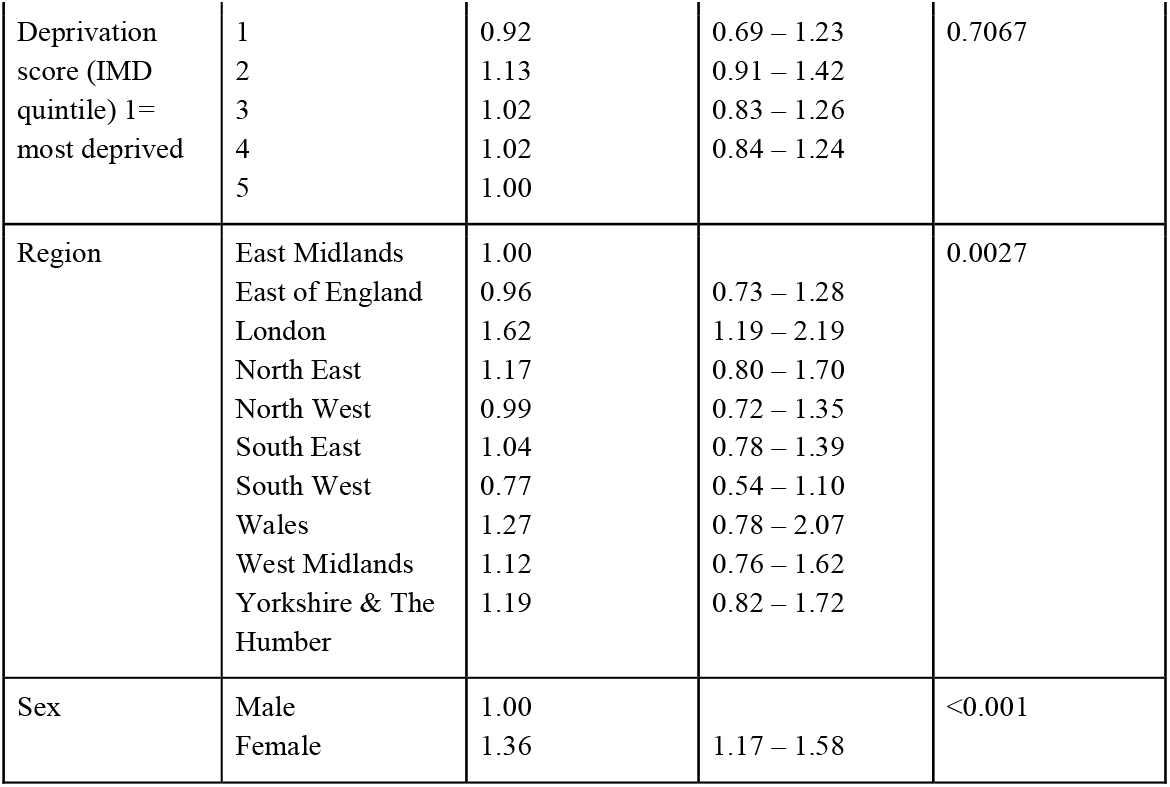
Multivariate analysis of number of contacts outside household, non household activities and risk of infection (all variables are mutually adjusted)

